# Demonstration of a Decongestant Effect of “Coldamaris Akut” Compared to Saline Nasal Spray in Participants Suffering from Seasonal Allergic Rhinitis

**DOI:** 10.1101/2024.05.03.24306805

**Authors:** Nicole Unger-Manhart, Martina Morokutti-Kurz, Petra Zieglmayer, Patrick Lemell, Markus Savli, René Zieglmayer, Hanna Dellago, Eva Prieschl-Grassauer

## Abstract

**Purpose:** Carrageenan-containing nasal sprays are known to alleviate symptoms of common cold and allergic symptoms by building a barrier against airborne intruders. The objective of this study was to develop a hyperosmolar nasal spray with barrier-forming properties and to demonstrate its decongestant effect in the context of allergic rhinitis.

**Methods:** The efficacy of the nasal spray components was first demonstrated *in vitro* by a virus replication inhibition, water absorption, and barrier assay. Clinical efficacy was assessed in a randomized, controlled, crossover trial, where adults with a history of severe seasonal allergic rhinitis were exposed to grass pollen allergens under controlled conditions for a total of 6 hours. Participants received either the carrageenan- and sorbitol containing nasal spray (CS) or saline solution (SS) after 1h45min of allergen exposure. After one week, participants repeated the exposure, receiving the treatment (CS or SS) they had not received before. The primary efficacy endpoint was the mean change in ’Nasal Congestion Symptom Score’ (NCSS) during the allergen exposure. Secondary efficacy endpoints were nasal airflow, nasal secretion, total nasal symptom score (TNSS), total ocular symptom score (TOSS) and total respiratory symptom score (TRSS).

**Results:** Preclinical assays showed virus-blocking, barrier building and water withdrawing properties of the CS components. In the clinical study, a total of 46 participants were screened, 41 were randomized and 39 completed the study. There was no significant difference in mean NCSS change from pre- to post-treatment between CS and SS (mean difference of 0.02 [95% CI -0.19; 0.24] during the first 2 hours after treatment) when analyzed by intention-to-treat. However, nasal airflow increased over time after treatment with CS, while it declined after SS, leading to a growing difference in airflow between CS- and SS-treated participants (p=0.039 at 6:00h). The anterior nasal airflow increased after treatment in 23/38 (61%) of the CS treated participants, compared to only 13/38 (34%) of the SS treated participants (p=0.024). The mean nasal secretion over 2-6 h was reduced by 1.00 g or -25% after CS (p=0.003) compared to pre-treatment, while it was reduced by only -0.50 g after SS (p=0.137). No significant differences in TNSS, TOSS and TRSS were observed between CS and SS treatments.

**Conclusion:** CS builds a barrier at the mucosa against viruses and dust and is safe and effective in alleviating nasal congestion, nasal airflow and nasal secretion in adults with grass pollen allergy.

**Trial registration:** NCT04532762

## Introduction

Nasal congestion, also described as fullness, blockage, or obstruction of the nasal cavity, is a frequently described symptom in clinical practice. It can significantly impair quality of life, reduce daytime productivity at work or school, and negatively impact night-time sleep time and quality.^1^ Nasal congestion is usually treated with local decongestants like Xylometazoline or Oxymetazoline. Unfortunately, rebound swelling of the mucosa is observed upon prolonged use of these topical vasoconstrictors. This often leads to a gradual overuse and a vicious circle of self-treatment, which patients are often not aware of.^2,3^

Nasal congestion is caused by air-borne irritants like tobacco smoke or dust, or by viruses and allergens which cause viral and allergic rhinitis and sinusitis, respectively. Allergic rhinitis is a type I allergic reaction where otherwise innocuous allergens such as pollen or animal dander crosslink receptor bound IgE on mast cells.^4^ This crosslinking results in a biphasic response. The early phase is characterized by the release of pre-formed mediators such as histamine which cause characteristic symptoms like pruritus, rhinorrhea, sneezing, and nasal congestion. The late phase is characterized by the release of newly synthesized mediators such as cytokines and chemokines. The latter strongly contribute to inflammation and thereby to a worsening of the disease. Seasonal allergic rhinitis or hay fever is caused by seasonal peaks in the airborne load of pollens and is the most common type of allergic rhinitis. It is one of the most common chronic conditions in high-income countries^5^ and it is estimated that in Europe, up to 40% of the population suffer from pollen allergy.^6,7^ In contrast to viral rhinitis, which is usually self-limiting with symptom duration of about 1 to 2 weeks, symptoms of allergic rhinitis can continue over longer periods. Allergic patients using topical decongestion are therefore at higher risk of the rebound effect and would benefit from a decongestant that does not induce this habituation effect.

Marinomed Biotech AG has developed nasal sprays based on iota-carrageenan (Carragelose^®^), a natural polymer from red seaweed, which forms a protective layer on mucosal surfaces that prevents viruses and allergens from interacting with the mucosal surface. Carragelose^®^ is certified for marketing in the EU, parts of Asia and Australia, as a component of nasal sprays, throat sprays and lozenges. Previous studies have shown that carrageenan-containing nasal sprays have a broad, non-specific mode of action and prevent attachments of small particles like virus or pollen to mucosal cells. This has been shown by us and others pre-clinically,^8–10^ and clinically.^11–17^ Carrageenan-containing nasal sprays reduce the symptoms of common cold and the viral load in nasal lavage.^14^ Symptom duration is shorter and viral titers in nasal fluids decrease faster in patients of common cold when treated with carrageenan-containing nasal spray compared to placebo.^12,13^ Since the virus-blocking effect of carrageenan is based on its physical barrier function, we hypothesized that it can act also against other small particles like pollen, resulting in the alleviation of AR symptoms.

To broaden the beneficial effect of our nasal spray, we wanted to add a decongestant activity by enhancing the osmolarity of the solution. This causes outflux of water from the nasal mucosa cells, thereby reducing mucosal swelling and hence nasal congestion. A hypertonic nasal spray containing carrageenan combines decongestant and anti-viral activity. Hypertonicity could be achieved by addition of ionic and/or non-ionic osmolarity givers like sodium chloride (NaCl). However, carrageenans change their conformation depending on the ionic strength of the environment.^18,19^ Enhancing osmolarity using NaCl might therefore affect their anti-viral properties. Alternatively, hypertonicity could be achieved by adding sorbitol, a water-soluble, membrane impermeant polyol (sugar alcohol) that is frequently used in food processing to preserve moisture and add sweetness and texture.

Here, we report preclinical in vitro and ex vivo data that are the basis for optimization of the decongestant nasal spray formulation. Furthermore, we show results of a randomized, controlled, crossover clinical trial on a decongestant effect of the CS in adults with a history of severe seasonal allergic rhinitis (SAR). The primary objective of this trial was to demonstrate a decongestant effect on the nasal mucosa of the CS in comparison with 0.5% saline solution nasal spray (SS). The secondary objective was to demonstrate the clinical performance of the CS in comparison with saline solution as assessed by objective measurements of nasal airflow and nasal secretion as well as patient-reported nasal, ocular and respiratory symptoms.

## Methods

### Preclinical assays

#### In vitro viral inhibition assay

To test if osmolarity could be adjusted with NaCl without compromising the virus-blocking effectiveness of carrageenan, a series of formulations containing 1.2 mg/ml iota-carrageenan and 0.4 mg/ml kappa-carrageenan with sodium chloride concentrations between 0.5% and 2.3% were tested against Human rhinoviruses HRV1a and HRV8. Hela cells were seeded in 96-well plates. 4-fold concentrated serial dilutions of the test sample (CS containing varying concentrations of NaCl) and 4-fold concentrated virus dilution were prepared. Equal volumes of virus and test sample dilutions were mixed and incubated at RT for 30 minutes. The mixture was diluted with an equal volume of medium with 4% fetal bovine serum and antibiotic/antimycotic before it was added to the cells for infection at a multiplicity of infection (MOI) of 0.7. After 48 hours at 33°C, cells were washed, and viability was assessed with Alamar Blue staining. Viability was corrected for toxicity of increasing salt concentrations and normalized to the viability of non-infected cells. The same experimental set-up was used to test viral inhibition effectiveness of the final formulation of the commercial product, containing 1.2 mg/ml iota-carrageenan, 0.4 mg/ml kappa-carrageenan, 0.5% NaCl, and 7% sorbitol in citrate/phosphate buffer. Half-maximal inhibitory concentrations (IC50) were calculated with XLfit Excel add-in version 5.3.1. Results were normalized to toxicity and non-infected control.

All percentages referring to nasal spray components here and in the following subsections are % weight/volume.

#### Hemagglutination assay

This assay was applied to assess anti-viral activity against coronavirus hCoV OC43. On a 96-well plate, two hemagglutination units of hCoV OC43 per well are incubated with a semi logarithmic dilution series of test or control samples for 10 min at RT (final concentrations: 0.002-3µg/ml iota-carrageenan diluted in 0.5% to 2.6% NaCl with or without 7% sorbitol and McIlvaine buffer). A suspension of chicken red blood cells (1% v/v in PBS) is added to each well to allow hemagglutination of RBCs by the virus for 1.5 hrs at 4°C. At the time point of assay evaluation, control RBCs in the absence of carrageenan are fully agglutinated by the virus, whereas inhibition of hemagglutination can be observed in samples treated with carrageenan up to a certain dilution factor. The minimal inhibitory concentration of each sample is noted for comparison of the anti-viral effectiveness of each sample under these assay conditions. As an internal control, a specific batch of iota-carrageenan is used (assay reference).

#### Ex vivo dehydration assay

The swine nasal mucosa was received from “University Clinic for Swine” at the University of Veterinarian Medicine Vienna. The nasal mucosa was excised from euthanized pigs and punched out into equal circular pieces with a diameter of 10mm. The mucosa pieces were weighed and put, the mucosa-site upward, into 48-well plates. 250 µl test solution was added to each well. Test solutions were iota- and kappa-carrageenan with 0.5% NaCl and 7% sorbitol; iota- and kappa-carrageenan with 0.5% NaCl without sorbitol; and a 2.4% NaCl solution. The plate was incubated for 60 minutes at 37°C, after which the mucosa pieces were weighed again.

#### In vitro barrier assay

A 1.25% agar solution was filled into the wells of a 96-deep-well plate and was left to solidify o/n at 4°C. 200 µl of CS and of negative control were added on top of the agar block. The negative control sample contained sorbitol and NaCl in same concentration as in CS but did not contain the barrier forming component carrageenan. Fluorescent beads of 0.3 µm or 1.0 µm, respectively, were added and incubated for 3h at RT. Following multiple wash steps with 0.5% NaCl solution, beads were extracted from agar blocks using 0.1% Tween20 in PBS o/n at 4°C with 900rpm shaking. Extraction supernatants were transferred into a 96-well black flat bottom plate and analyzed in a plate photometer with an excitation and emission wavelength of 485nm and 520nm, respectively. Percent blocking was calculated relative to the amount of beads extracted from the negative control.

### Clinical study

#### Study design

This was a prospective, controlled, double-blinded randomized two-way cross-over single site study in adult female and male participants with severe grass pollen induced seasonal allergic rhinitis (SAR). The study evaluated two treatments, namely the carrageenan- and sorbitol containing nasal spray (CS) and a saline solution (SS) nasal spray. The study was conducted at the Vienna Challenge Chamber (VCC) in Vienna, Austria. The Ethics Committee of the City of Vienna oversaw trial conduct and documentation. The study was designed to include 5 visits. At visit 1 (screening visit), participants were screened for appropriate allergic response. At visit 2, which could be done on the same day as visit 1, medical and allergic history and inclusion/exclusion criteria were assessed and blood samples for safety lab were withdrawn. At visit 3, scheduled 7 days after visit 2, participants were randomized to one of the two treatment arms (CS or SS) in a fully blinded fashion (details of randomization see below) and underwent their first six-hour allergen challenge session. Approximately 1hour and 45 minutes after start of allergen exposure, participants were dosed with the treatment they had been randomized to, and continued exposure for a total of 6 hours. (first treatment block). At visit 4, scheduled 7 days after visit 3 to allow complete symptom relief from the previous challenge, participants were exposed to the second allergen challenge (second treatment block) and crossed over to the treatment that they had not received in the first block. A follow-up visit (end of study visit, visit 5) was scheduled one week after the second treatment block. Participants were asked to record AEs and the use of concomitant medications for the entire duration of the trial.

#### Participants

Participants were female and male adults aged between 18 and 65 years of any ethnicity/race, with a documented clinically relevant allergic history of moderate to severe SAR to grass pollen for the previous two years. Participants were selected from the VCC database and had to satisfy all inclusion and exclusion criteria to be enrolled into the study. Key inclusion criterion was a moderate to severe response to standard grass pollen allergen mixture within the first 2 hours in the VCC, defined as total nasal symptom score (TNSS) of at least 6 (out of 12) with the necessity to score at least “moderate = 2” for the single symptom ‘nasal congestion’. TNSS is the sum of ‘nasal congestion’, ‘rhinorrhea’, ‘itchy nose’ and ‘sneezing’, each scored on a categorical scale from 0 to 3. In addition, participants had to fulfill the following inclusion criteria: a positive Skin Prick Test (SPT) response (wheal diameter at least 3 mm larger than diluent control) to grass pollen solutions (standard Allergopharma) at screening or within the last 12 months prior to study start; positive serum specific IgE against recombinant major allergen components of the used grass pollen e.g., g6 (specific CAP IgE ≥0.70 kU/L) at screening or within the last 12 months prior to study start; and a forced expiratory volume in 1 second (FEV1) of at least 80% of reference value^20^ at screening. Asthma patients were allowed into the study only if the asthma condition was mild or intermittent, and if not treated with steroids. Exclusion criteria comprised prior and ongoing conditions, diseases and treatments that may interfere with the study intervention and outcomes. Female participants of child-bearing potential were required to use birth control.

#### Randomization and blinding

Randomization numbers were allocated to the study participants in ascending order of their Screening Numbers following their attendance at Visit 3 (first treatment block). They were randomized using a cross-over randomization with balanced blocks. All personnel involved in the study, including investigators, site personnel, and sponsor’s staff were blinded to the randomization codes. Persons responsible for labeling of investigational products were un-blinded, but not involved in other study activities. Un-blinding occurred at the end of the study.

#### Interventions and procedures

During each treatment period, participants were exposed to standard grass pollen allergen mixture in the VCC for six hours using a validated method.^21,22^ During the challenge session, participants were under constant supervision by, and could communicate with, medical staff outside the chamber. The chamber was charged with 100% fresh air, which was conditioned (filtered, heated, dried, cooled, and humidified) and then loaded with the challenge agent, a mixture of four grass pollen species (Timothy, Orchard, Perennial rye and Sweet vernal grass) (Allergon SB, Sweden). Air temperature (24°C), humidity (40%) and allergen load (1500 grains/m^3^) were constantly monitored and maintained. During the 6 hours challenge, subjective nasal symptoms (nasal congestion, rhinorrhea, itching, sneezing) as well as ocular and respiratory symptoms were recorded every 15 minutes. Nasal airflow was measured by active anterior rhinomanometry (AAR) at a pressure difference of 150 Pascal across the nasal passages (sum of the right and left nostril values). Nasal airflow was evaluated immediately before and every 30 minutes during exposure, with an additional assessment at timepoint 2h 15min. Nasal secretion was evaluated by weighing paper tissues used by the participants during their stay in the chamber and collected every 30 minutes. 1h 45min after entering the challenge chamber, i.e., after developing pronounced allergic nasal symptoms including nasal congestion, participants applied 1 puff per nostril of either CS or SS. This resulted in a residual observation period of 4h 15min.

CS contained 1.2mg/ml iota-carrageenan, 0.4 mg/ml kappa-carrageenan, 7% (w/v) sorbitol, 0.5% (w/v) sodium chloride, 1 mg/ml ethylene diamine tetra acetate, buffer and purified water. SS contained 0.5% sodium chloride in water.

#### Endpoints

The primary efficacy endpoint was the mean difference between CS and SS of the ’Nasal Congestion Symptom Score’ (NCSS) measured every 15 min during allergen exposure. Secondary efficacy endpoints were nasal airflow as assessed by active anterior rhinomanometry, total nasal symptom score (TNSS; sum of the symptoms ‘nasal congestion’, ‘rhinorrhea’, ‘itchy nose’, and ‘sneezing’), total ocular symptom score (TOSS; sum of the symptoms ‘ocular itching’, ‘redness’, ‘watery eyes’), total respiratory symptom score (TRSS; sum of the symptoms ‘cough’, ‘wheeze’, ‘dyspnea’), and nasal secretion. Each individual symptom of NCSS, TNSS, TOSS and TRSS was rated on a scale from 0 to 3, whereas “0” corresponded to “no symptoms”, “1” to “mild symptoms” (easy to tolerate), “2” to “moderate symptoms” (bothersome, but tolerable) and “3” to “severe symptoms” (hard to tolerate). Safety assessments included frequency and severity of AE, related AE and serious AE (SAE) throughout the study. In addition, vital signs (blood pressure, pulse rate, temperature and breathing frequency) were assessed at every visit, pre-and post-challenge. Lung function was assessed at screening as well as before allergen challenge and every 2 hours during the allergen challenge by measuring the Forced Expiratory Volume in 1 second (FEV1) using a Piston Spirometer. Physical examination, laboratory blood analysis and ECG were conducted at screening and at the follow-up visit.

#### Statistical analysis

Sample size calculation was based on the expectation of a mean difference of 0.6 points with standard deviation of 1.1 (SS = 2, CS = 1.4, effect size d=0.56 and a power = 90%) which was derived from previous studies. Thus, n=36 participants were needed at an alpha level of p=0.05. Considering the dropout rate of 10-15%, up to 50 participants needed to be screened to randomize about 42 participants in order to get evaluable data from at least 36 participants.

Safety analyses including vital signs, laboratory data and AEs, were carried out in the safety population defined as all participants starting the challenge provocation qualification session.

Efficacy was analyzed in the Full Analysis Set (FAS) and in the Per-Protocol Set (PPS). The FAS comprised all participants who were randomized and was analyzed following the intent-to-treat (ITT) principle, according to the treatment they have been assigned at randomization. The PPS comprises all participants in the FAS who did not have any clinically important protocol deviation. The primary efficacy variable was analyzed in a confirmatory way between the two conditions CS and SS, assuming superiority for CS versus SS. The null hypothesis was defined as:

*Mean NCSS [Delta pre-treatment (1:45h) - post-treatment (mean 2-4h)] {*CS*} ≤ Mean NCSS [Delta pre-treatment (1:45h) - post-treatment (mean 2-4h)] {SS}*

The alternative hypothesis was formally defined as:

*Mean NCSS [Delta pre-treatment (1:45h) - post-treatment (mean 2-4h)] {*CS*} > Mean NCSS [Delta pre-treatment (1:45h) - post-treatment (mean 2-4h)] {SS}*

A 95% confidence interval for the mean difference of the two treatments was calculated. The superiority comparison of CS versus SS was performed using analysis of variance (ANOVA) appropriate for the cross-over design. Period (first or second treatment block) was included in the analysis model as a fixed effect to confirm the assumption of no period effect. Participant was included in the model as a random effect. Superiority was to be postulated if the lower bound of the 95% confidence interval was >0.

Secondary efficacy variables were analyzed in an explorative sense and are presented using descriptive methods. Exploratory efficacy analysis was performed for mean differences between the two treatments for consecutive intervals from 2h onward to 6h analogous to the primary efficacy analysis. Respective statistical tests and p-values are to be regarded as descriptive and not as tests of hypotheses.

All attempts were made to collect all data per protocol. Missing or invalid data was neither replaced nor extrapolated. Outliers were not excluded from the primary analyses. Significance level was set to alpha=5%. R version 4.0.3 was used for all statistical analyses.

## Results

### Results Part 1: Preclinical Development

Carrageenan containing nasal sprays are used to prevent and treat viral infections of the respiratory tract by blocking the viruseś attachment to the mucosa. To enhance the benefit and broaden the applicability of the barrier-forming nasal spray, a decongestant effect should be added to the formulation. Usually, intranasally applied hyperosmotic saline solutions are used to withdraw water from the nasal mucosa, thereby reducing intranasal swelling. However, we found that increasing salt concentrations reduced the carrageenan’s capacity to block the attachment of human rhinovirus and of human coronavirus to cells. As shown by IC50 values in **Table 1**, increasing sodium chloride concentrations reduced the virus-blocking capacity of the carrageenan against human rhinovirus HRV1 and HRV8 as well as against Coronavirus hCoV OC43 in a dose-dependent manner. Therefore, the formulation was adjusted to 0.5% sodium chloride to preserve the carrageenan’s beneficial virus-blocking effect. To achieve hyperosmotic activity, sorbitol was added to the formulation at a concentration of 7%, which increased the formulatiońs osmolarity, but in contrast to high concentrations of sodium chloride, preserved the virus-blocking activity of carrageenan (also shown in **Table 1**).

**Table 1:** In vitro data: Virus-blocking effectiveness against HRV1a, HRV8 and hCoV OC43. Minimal inhibitory concentration of the various formulations determined in a virus inhibition assay (for HRV1 and HRV8) or a hemagglutination inhibition assay (for hCoV OC43).

| Effectiveness of various formulations | IC<sub>50</sub><br>[µg/ml] | IC<sub>50</sub> 95% CI<br>[µg/ml] |
| --- | --- | --- |
| HRV1a |  |  |
| Carrageenan + 0.5% NaCl | 1.8 | 0.7; 3.0 |
| Carrageenan + 0.9% NaCl | 5.6 | 4.0; 7.1 |
| Carrageenan + 2% NaCl | 26.5 | 23.0; 30.0 |
| Carrageenan + 2.3% NaCl | 40.7 | 35.0; 46.6 |
| Fold Change (Carrageenan + 0.5% NaCl) vs. (Carrageenan + 2.3% NaCl) | 22.3 |  |
| Carrageenan + 0.5% NaCl + 7% Sorbitol | 3,7 | 2.2; 5.3 |
| Carrageenan + 2.3% NaCl | 104,5 | 82.8; 126.2 |
| Fold Change (Carrageenan + 0.5% NaCl + 7% Sorbitol) vs. (Carrageenan + 2.3% NaCl) | 27.9 |  |
| HRV8 |  |  |
| Carrageenan + 0.5% NaCl | 2.3 | 1.0; 3.6 |
| Carrageenan + 0.9% NaCl | 4.1 | 2.9; 5.3 |
| Carrageenan + 2% NaCl | 8.1 | 5.5; 10.7 |
| Carrageenan + 2.3% NaCl | 15.6 | 8.8; 22.4 |
| Fold Change (Carrageenan + 0.5% NaCl) vs. (Carrageenan + 2.3% NaCl) | 6.9 |  |
| Carrageenan + Buffer + 0.5% NaCl + Sorbitol | 0.8 | 0.7; 1.0 |
| Carrageenan + 2.3% NaCl | 2.3 | 1.5; 3.1 |
| Fold Change (Carrageenan + 0.5% NaCl + 7% Sorbitol) vs. (Carrageenan + 2.3% NaCl) | 2.9 |  |
| hCoV OC43 |  |  |
| Carrageenan + 0.5% NaCl | 0.007 | n.a. |
| Carrageenan + 0.9% NaCl | 0.007 | n.a. |
| Carrageenan + 2.0% NaCl | 0.080 | n.a. |
| Carrageenan + 2.3% NaCl | 0.080 | n.a. |
| Carrageenan + 0.5% NaCl + 7% sorbitol | 0.007 | n.a. |
| Fold Change (Carrageenan + 0.5% NaCl) vs. (Carrageenan + 2.3% NaCl) | 11.4 |  |
| Fold Change (Carrageenan + 0.5% NaCl + 7% Sorbitol) vs. (Carrageenan + 2.3% NaCl) | 11.4 |  |

Note: Bold borders mark individual experiments.
Abbreviations: IC<sub>50</sub>, inhibitory concentration neutralizing 50% of the virus; CI, confidence interval; NaCl, sodium chloride;
Carrageenan, 1.2 mg/ml iota-carrageenan and 0.4 mg/ml iota-carrageenan; n.a., not applicable.

After confirming that addition of buffer did not influence the antiviral activity of carrageenan (data not shown), the final product was formulated with 1.2 mg/ml iota-carrageenan, 0.4 mg/ml kappa-carrageenan, 0.5% NaCl, 7% sorbitol in citrate/phosphate buffer with an osmolality of 787 mosmol/kg, corresponding to the osmolality of hyperosmolar saline solutions with concentrations of 2.3-3%. This formulation was then used for ex vivo experiments as well as for the clinical study. Ex vivo experiments showed that incubation of nasal porcine mucosa with CS or a 2.4% saline solution of similar osmolality withdrew considerable amounts of liquid from the mucosa, resulting in a weight loss of 21±5% and 14±8%, respectively. In comparison, the weight of the mucosa incubated with carrageenan in 0.5% NaCl remained equal (weight change of 1±6%), indicating that the hyperosmolality alone, and not the carrageenan, is responsible for the weight loss (**Figure 1**). These results demonstrate a beneficial effect of sorbitol when added to the CS that could support nasal decongestion via its water draining properties.

**Figure 1:**
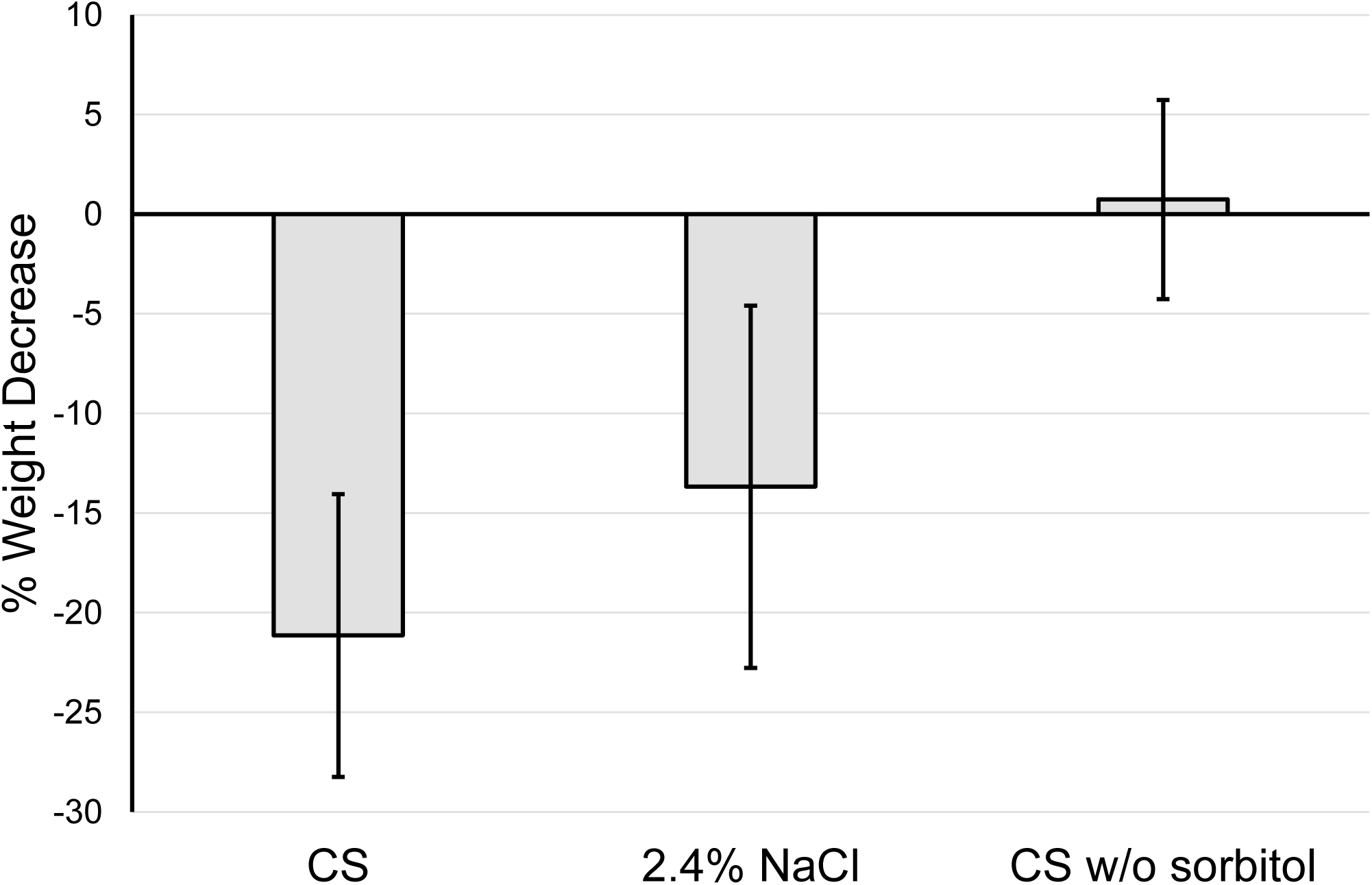
Ex vivo assay: Hyperosmolar effect of CS nasal spray with and without sorbitol. Weight decrease of ex-vivo porcine nasal mucosa after incubation for 60 minutes at 37°C in CS (carrageenan + 0.5% NaCl + 7% sorbitol in buffered aqueous solution), a 2.4% sodium chloride solution, or carrageenan + 0.5% NaCl in buffered aqueous solution without sorbitol (CS w/o sorbitol). Error bars represent standard deviation of replicates.

A proof of principle for the barrier function of carrageenan in the formulation containing 7% sorbitol and 0.5% NaCl was demonstrated by an in vitro barrier assay. This assay tests the ability of a sample solution to inhibit diffusion of fluorescent beads, serving as surrogate for particulate matter, into an agar block. As shown in **Figure 2**, CS nasal spray exhibited a blocking activity of 99±0% for beads of 0.3 µm diameter, and of 80±2% for beads of 1.0 µm diameter. This means that the protective layer formed by carrageenan allowed only 1% and 20%, respectively, of beads to reach the agar block, compared to the negative control. This indicates that the nasal spray can provide protection against external particles that might trigger or worsen allergic reactions.

**Figure 2:**
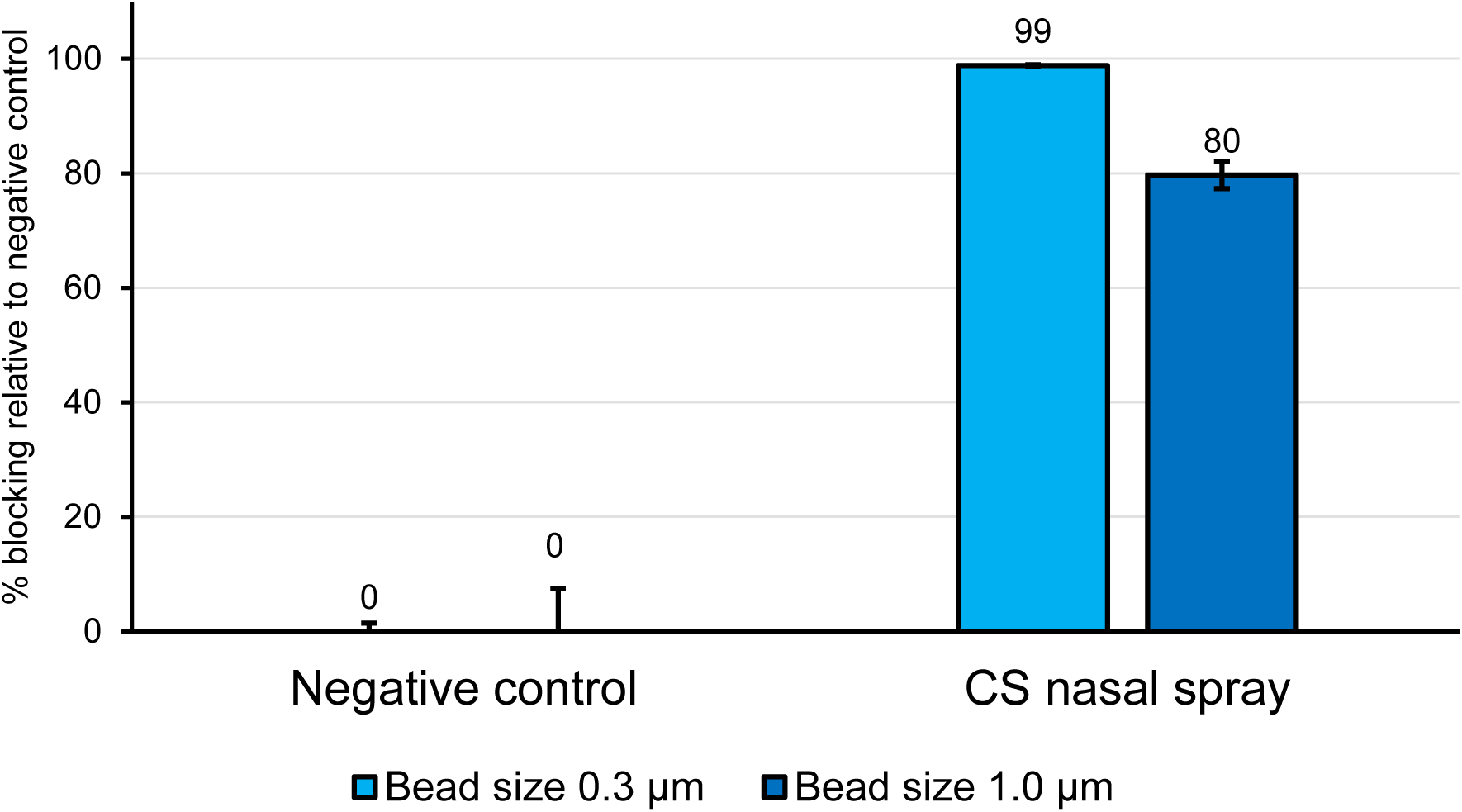
In vitro assay: Barrier function of CS nasal spray. Results of the percentage blocking activity of CS nasal spray relative to negative control (contains sorbitol and NaCl in same concentration as in CS but does not contain the barrier forming component carrageenan). Amounts of barrier-crossing beads were analyzed 180 minutes after application of beads. Cyan = % blocking activity for bead size of 0.3 µm; blue = % blocking activity for bead size of 1.0 µm. Error bars represent standard deviation of replicates.

### Results Part 2: Clinical Study

The potential of the CS to treat nasal congestion in humans was examined in a clinical study in patients with allergic rhinitis. **Figure 3, Panel A** gives a graphical overview of the study, **Panel B** depicts the assessment carried out during each treatment block. Between September and October 2020, a total of 46 participants were screened after giving informed consent and were included in the safety population. Of these, 41 participants fulfilled all in/exclusion criteria, were randomized to one of the two possible treatment sequences, and hence constitute the FAS. 2 participants discontinued, and 4 participants did not respond to either treatment with CS or SS and were thus excluded from the per PPS based on the finding that hypertonic saline nasal spray has no effect on nasal congestion in approximately 30% of the population.^23^ No other exclusionary protocol deviations were reported. **Figure 4** shows the flow of participants through the study.

**Figure 3:**
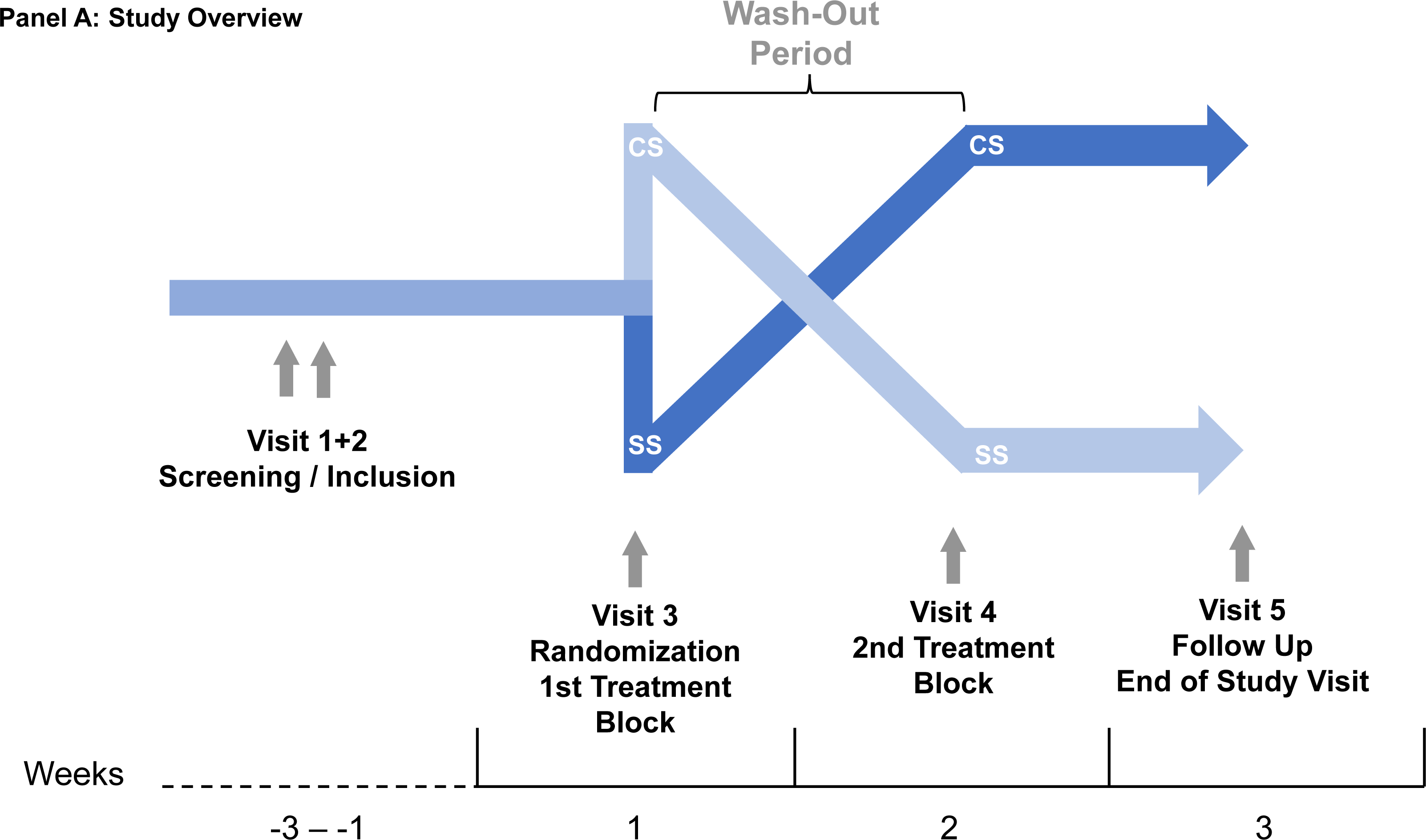
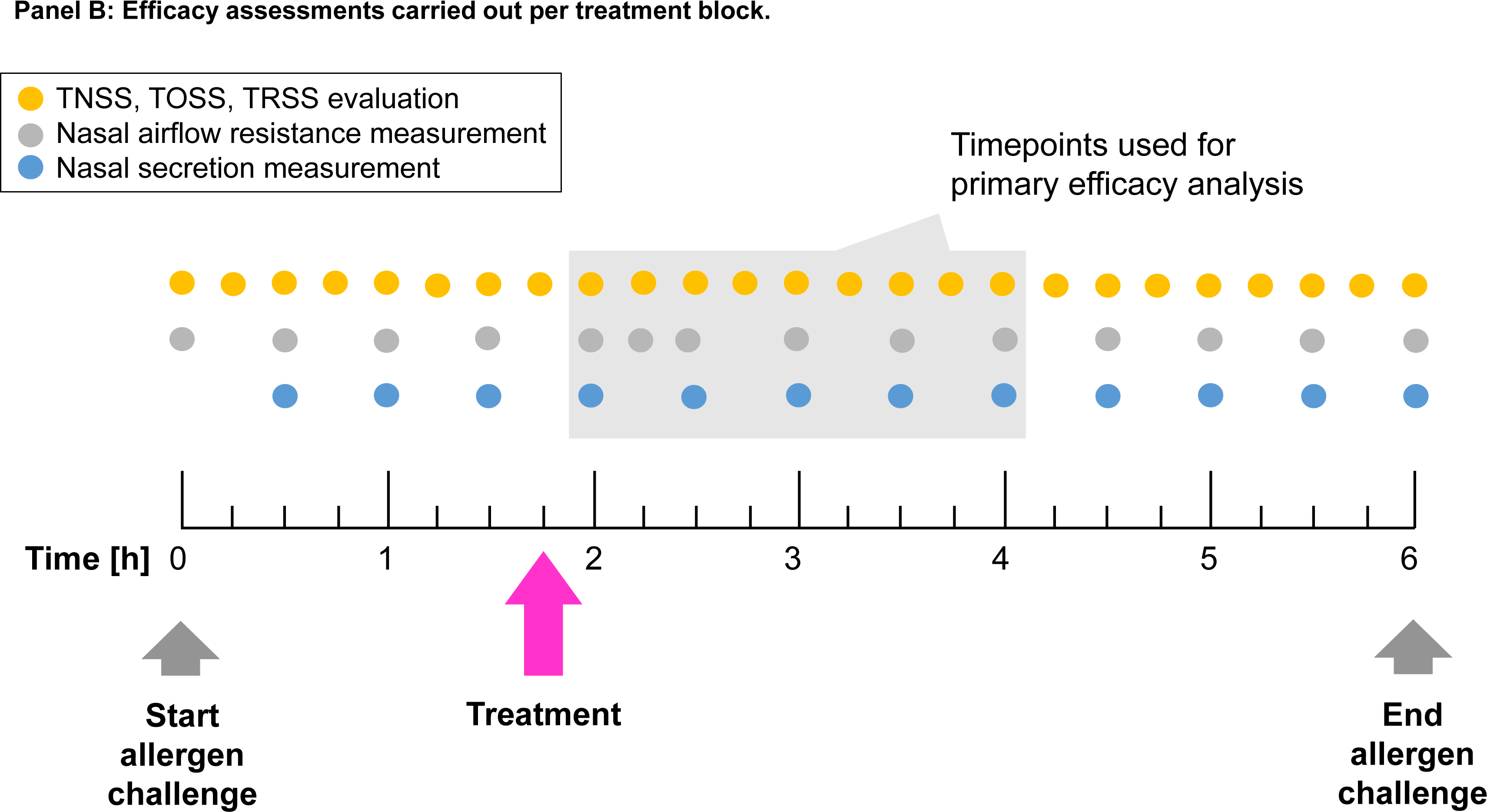
Clinical Study: Graphical Abstract.

**Figure 4:**
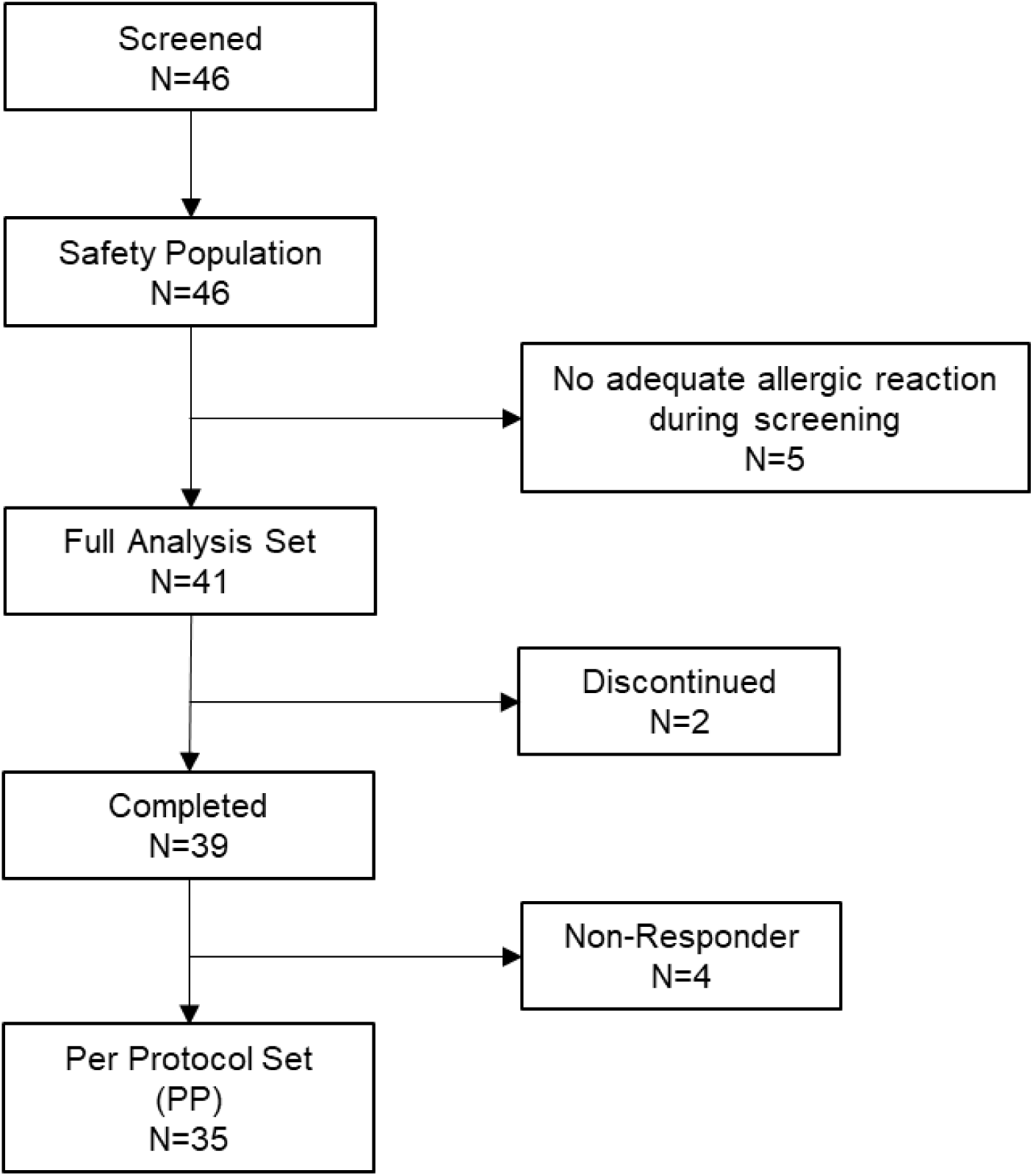
Clinical Study: CONSORT Flow Chart.

Demographic characteristics are summarized in **Table 2**. 27/46 (59%) of the participants were females, 19/46 (41%) were males. Participants were aged between 21 and 62 years, with a mean age of 34.6 years (SD 10.9). The mean BMI was 23.9 kg/m^2^, all participants’ BMIs were below 30, i.e., none of the participants was obese. All participants had a history of moderate to severe seasonal allergic rhinitis (SAR) to grass pollen with a prior duration of between 8 and 43 years, on average 23.5 years.

**Table 2:**
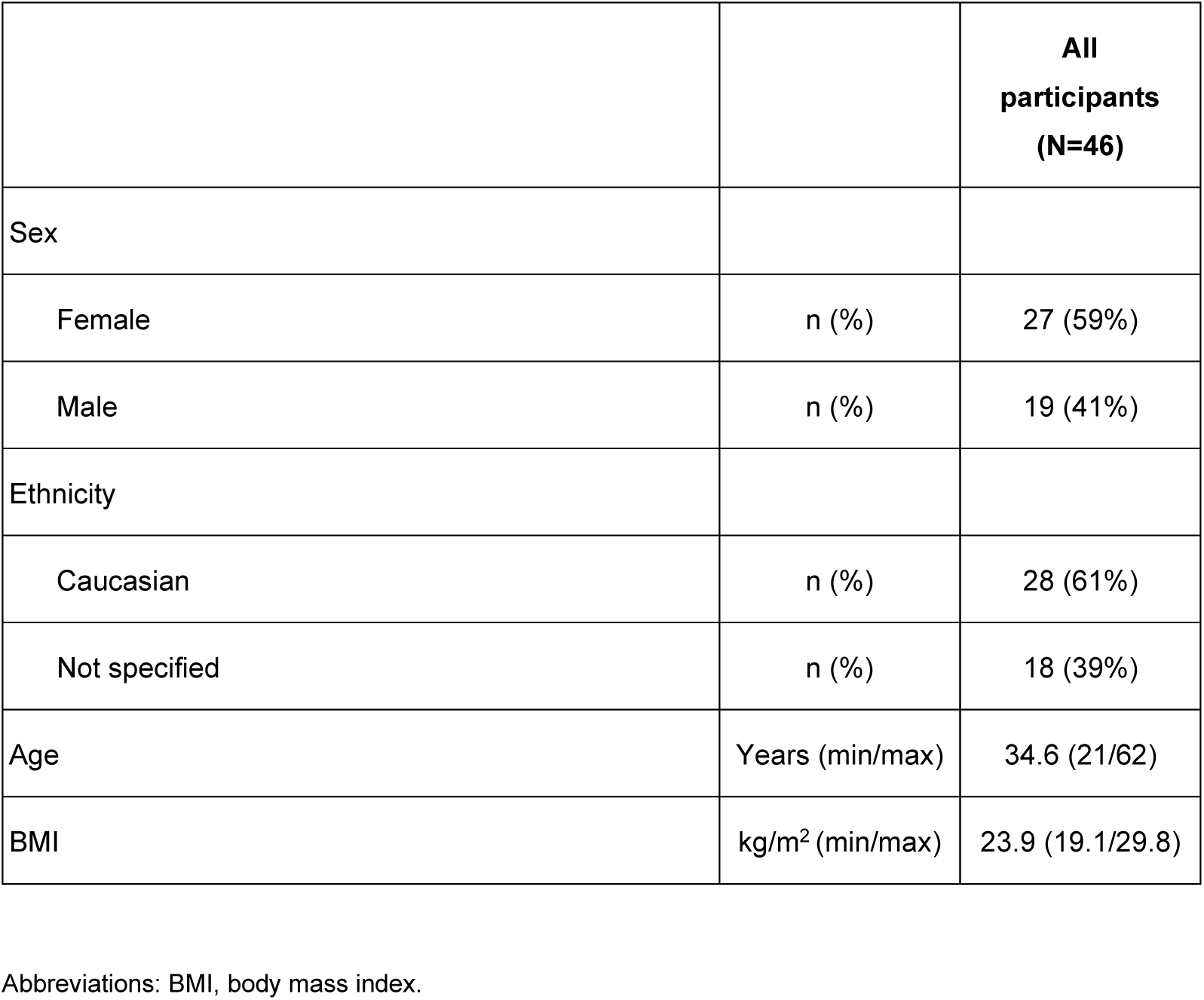
Clinical data: Demographic characteristics at baseline (Safety Population)

In the following, all efficacy results are shown for the FAS, analyzed by ITT. Results for the PP were similar as for the FAS.

All participants developed nasal congestion upon the start of the allergen challenge. The mean NCSS increased notably already after 15 min, further increased until timepoint 1h 45min, and was reduced upon intake of either CS or SS (**Figure 5A**). The overall mean NCSS was 0.1 (SD 0.3) before starting the allergen challenge (timepoint 00:00) and it increased to 2.3 (SD 0.7) after CS treatment group and 2.2 (SD 0.5) after saline solution treatment at timepoint 1h45min (**Supplementary Table S1**). However, only a small difference of 0.16 (SD 0.50) for CS and 0.11 (SD 0.53) for SS between pre-treatment NCSS (timepoint 1h45min, i.e., directly before the treatment), and the mean NCSS across the time interval 2-4h could be detected (**Supplementary Table S2**). No phase-effect (p-value >0.05, Wilcoxon test) and no carry-over effect (p-value >0.05, ANOVA) was observed. The mean difference between CS [Pre-treatment - ø(2-4h)] and SS [Pre- treatment - ø(2-4h)] across all participants was 0.02, 95% CI [-0.19;0.24], p >0.05 (paired t-test) (**Figure 5B**). With the lower bound of the 95% confidence interval <0, superiority of CS versus SS in terms of NCSS could not be established.

**Figure 5:**
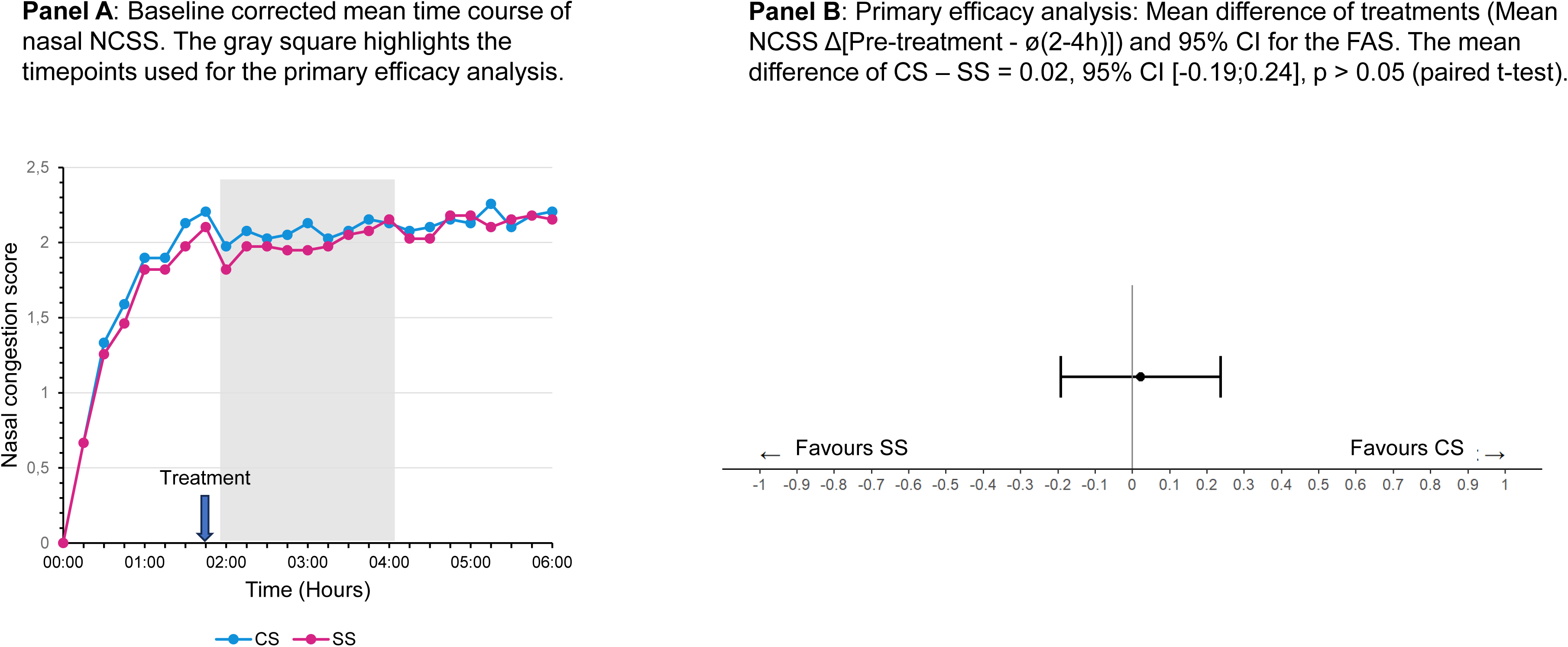
Clinical data: Nasal Congestion Symptom Score (NCSS) pre- and post-treatment during the grass pollen allergen exposure challenge for the FAS.

**Figure 6** shows the absolute nasal airflow in both treatments before treatment (timepoint 1h45min) and at the end of the allergen challenge period. In total, an increased anterior nasal airflow was measured in 23/38 (61%) of the participants after treatment with the CS, but in only 13/38 participants (34%) after SS treatment (**Table 3**). This difference between treatments was statistically significant (p=0.024, McNemar’s test for paired nominal data).

**Figure 6:**
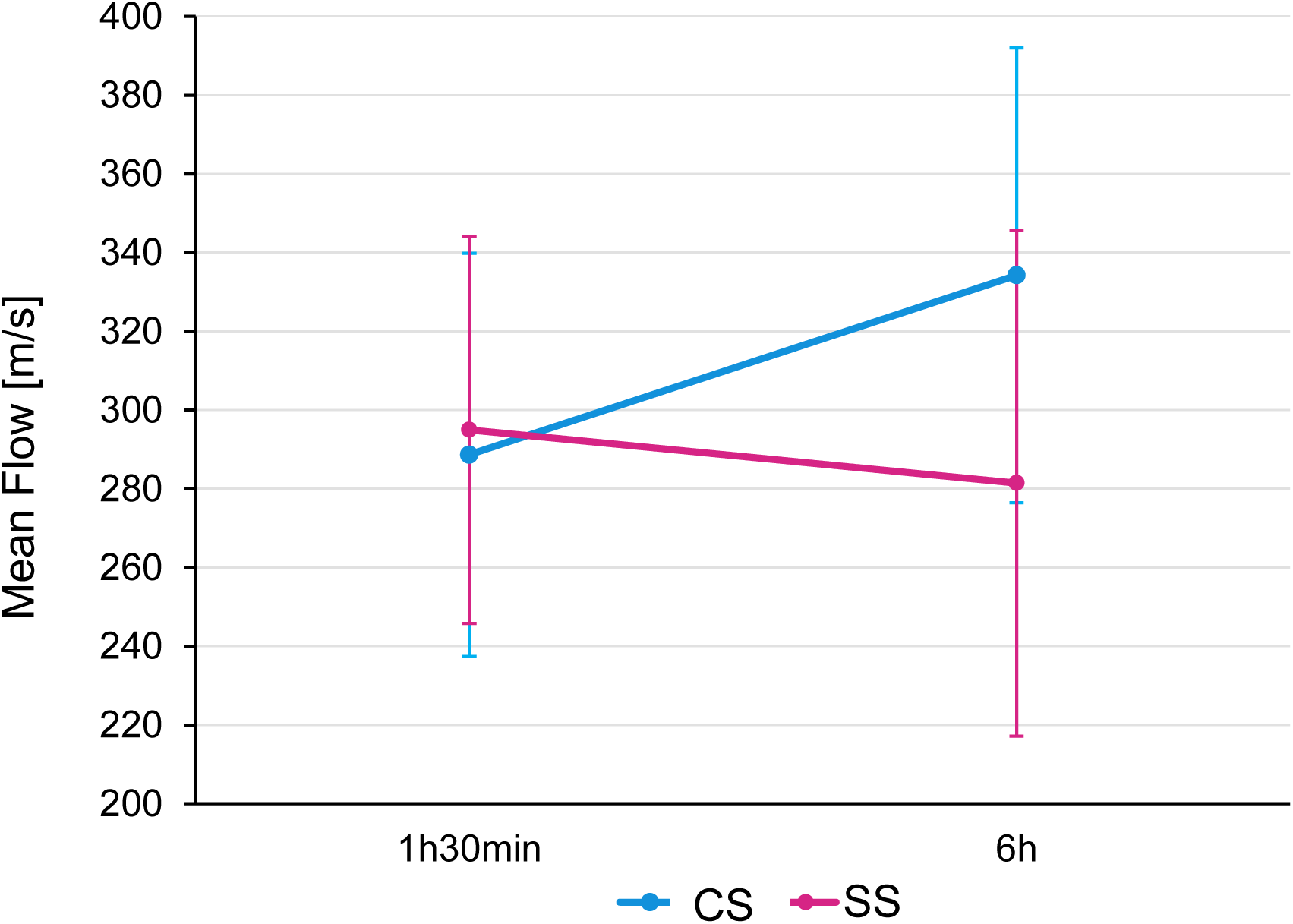
Clinical data: Anterior nasal airflow before and after treatment for the FAS. Mean airflow at timepoints 1h30min (before treatment) and 6h after start of allergen challenge. Error bars denote 95% CI. P=0.039 for comparison between treatments in difference from pre-treatment to timepoint 360 min.

**Table 3:** Clinical data: Improvement/worsening of airflow after 6h compared to pre-treatment (1h30min), evaluated within treatment groups for the FAS.

|  |  | CS (360 min - 90 min) |  |
| --- | --- | --- | --- |
|  |  | better or equal | worse |
| SS<br>(360 min - 90 min) | better or equal | 10 | 3 |
|  | worse | 13 | 12 |

Abbreviations: CS, carrageenan-sorbitol containing nasal spray. SS, saline solution.
p-Value: 0.024 (McNemar's test for paired nominal data for comparison between treatments)

In order to unravel the temporal dynamics that led to the post-treatment differences, we also followed nasal airflow changes over time by subtracting the mean pre-treatment value (timepoint 1h30min) from the mean post-treatment value of varying post-treatment periods (mean over 2-6h, 2:15-6h, 2:30-6h etc.). Positive values indicate higher nasal airflow post-treatment compared to pre-treatment. As shown in **Supplementary Figure S1**, treatment with the CS led to an increase of nasal airflow over the course of the 4 hours residual observation time compared to pre-treatment, while it declined in the SS group. This led to a significantly higher airflow in the CS group compared to the SS group at the end of the 6 hours treatment block: The difference between CS and SS in nasal airflow change from pre-treatment to the end of the 6h treatment block in the FAS (ITT) population was 54.29 ml/s (95% CI 2.92; 105.66). The difference was significantly in favor of the CS (p=0.04, paired t-test) (**Supplementary Table S3**).

Changes in nasal secretion from pre- to post-treatment were calculated in an analogous manner. In both groups, nasal secretion declined post-treatment when compared to pre-treatment. The difference in nasal secretion from pre- to post-treatment was more pronounced in the CS group than in the SS group (Figure 7). For the CS, the weight of nasal secretion changed from 3.99 g at pre-treatment to 2.99 g averaged over the entire residual observation time (2-6h), representing a mean tissue weight difference of -1.00 g or -25% (p=0.003, t-Test). After SS, the mean tissue weight difference from pre-treatment to 2-6h, was only -0.50 g (p=0.137, Wilcoxon signed rank test). These results indicate that nasal secretion declined more strongly after CS than after SS treatment (**Table 4 and** Figure 7).

**Figure 7:**
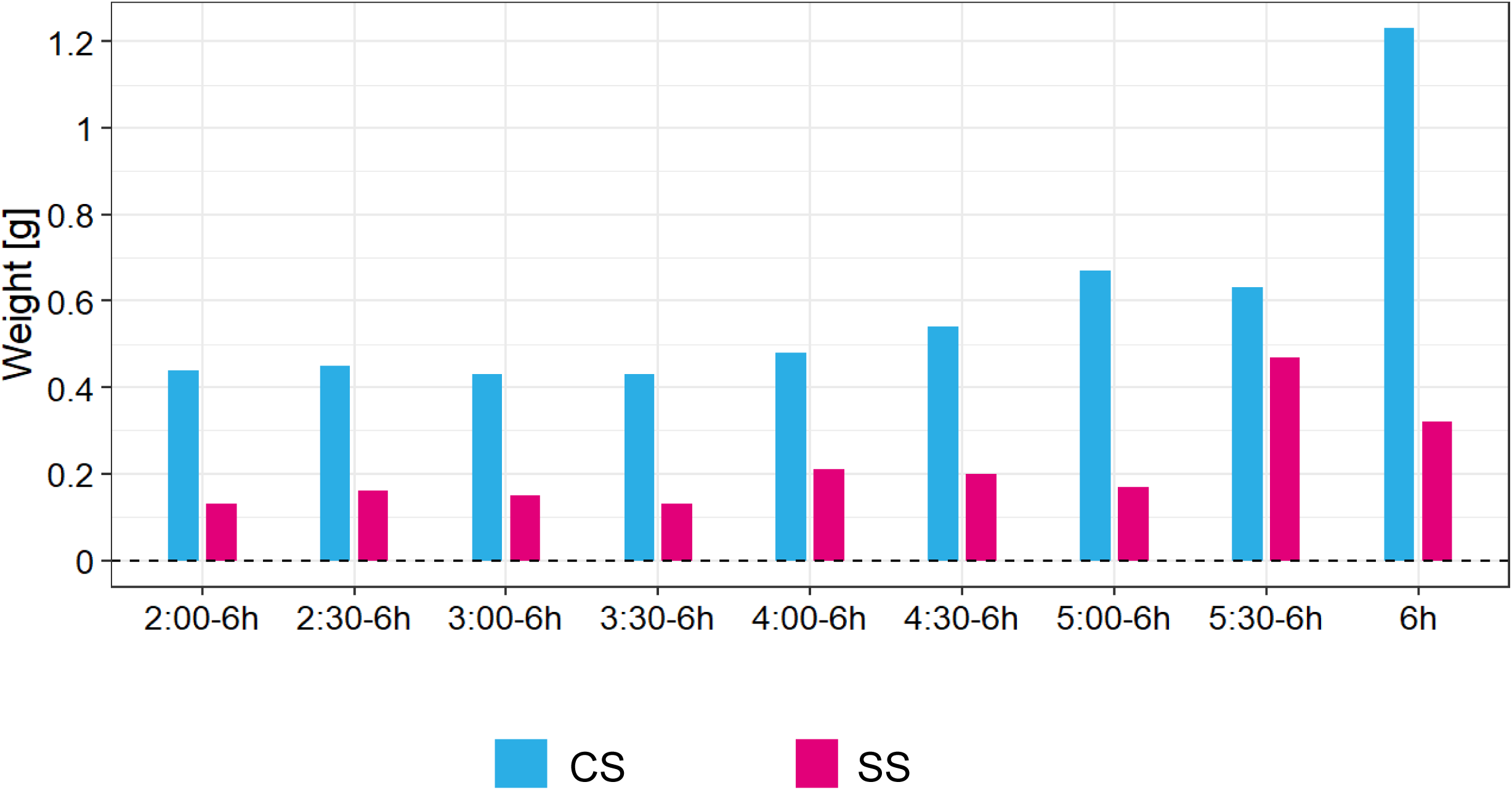
Clinical data: Median nasal secretion absolute differences to pre-treatment for the FAS. Differences in post-treatment nasal secretion compared to ptre-treatment after CS treatment (cyan) and saline treatment (magenta). Positive values indicate lower nasal secretion post-treatment compared to pre-treatment.

**Table 4:**
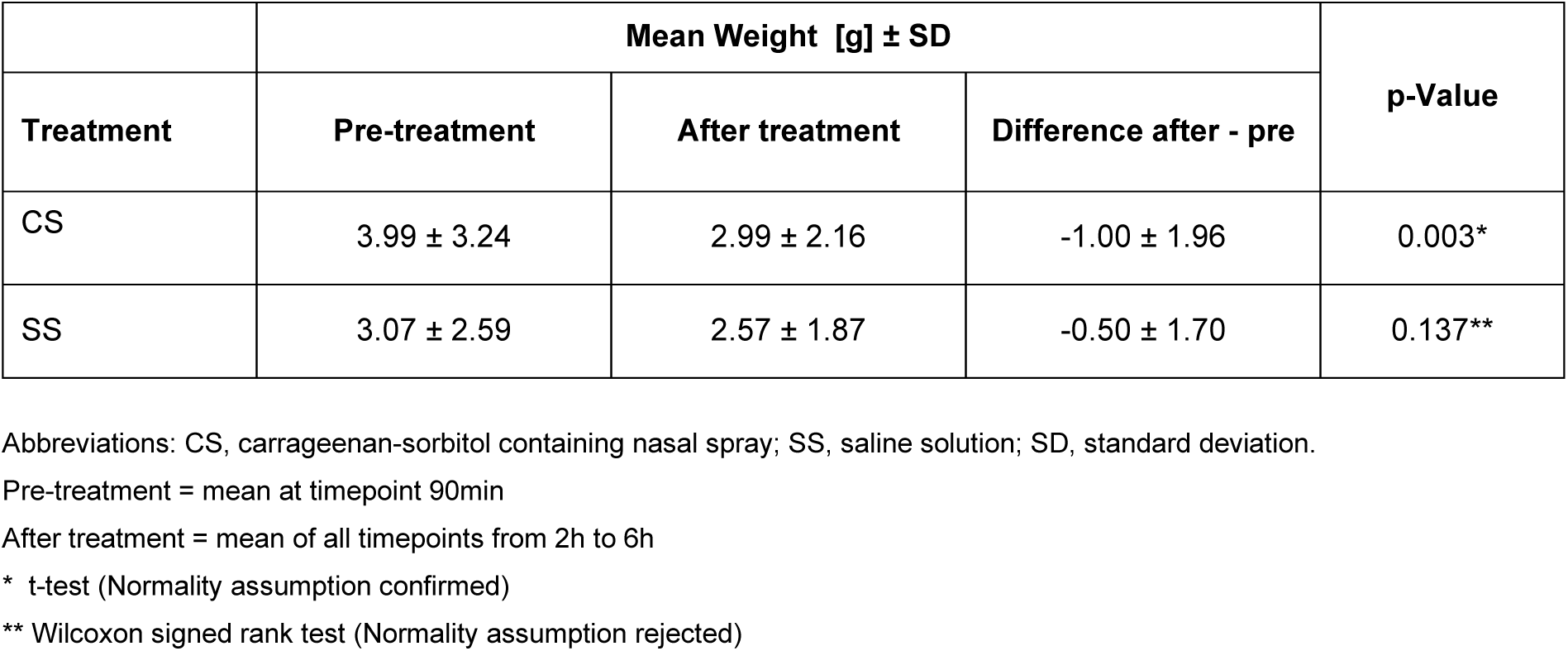
Clinical data: Tissue weight differences between pre-treatment [90 min] and the mean of all post-treatment timepoints [120-360 min] for the FAS.

| Treatment | Mean Weight [g] ± SD |  |  | p-Value |
| --- | --- | --- | --- | --- |
|  | Pre-treatment | After treatment | Difference after - pre |  |
| CS | 3.99 ± 3.24 | 2.99 ± 2.16 | -1.00 ± 1.96 | 0.003* |
| SS | 3.07 ± 2.59 | 2.57 ± 1.87 | -0.50 ± 1.70 | 0.137** |

Abbreviations: CS, carrageenan-sorbitol containing nasal spray; SS, saline solution; SD, standard deviation.
Pre-treatment = mean at timepoint 90min
After treatment = mean of all timepoints from 2h to 6h
\* t-test (Normality assumption confirmed)
\*\* Wilcoxon signed rank test (Normality assumption rejected)

TNSS, TOSS and TRSS over the 6 hours treatment block did not show any pronounced differences between CS and S group (data not shown).

In the safety population, a total of 3 adverse events occurred in 2 participants during the trial: pyrexia (mild), nasopharyngitis (moderate) and pharyngitis (severe) (**Table 5**). Pharyngitis and pyrexia occurred in the same participants 4 days after the first treatment block with SS. Nasopharyngitis occurred 4 days after the first treatment block with CS. None of them was considered related to the study treatment, none was serious, all were resolved by study end. Both participants missed the second treatment block and terminated the trial prematurely due to these AEs.

**Table 5:** Clinical data: Adverse events by SOC/PT and severity for the Safety Population (N=46).

| SOC | PT | Mild | Moderate | Severe | Total |
| --- | --- | --- | --- | --- | --- |
| **General disorders and administration site conditions** |  | **1** | **0** | **0** | **1** |
|  | Pyrexia | 1 | 0 | 0 | 1 |
| **Infections and infestations** |  | **0** | **1** | **1** | **2** |
|  | Nasopharyngitis | 0 | 1 | 0 | 1 |
|  | Pharyngitis | 0 | 0 | 1 | 1 |

Abbreviations: SOC, system organ class; PT, preferred term.

All vital signs and laboratory values showed no particular differences between baseline and follow-up visit (data not shown), indicating good tolerability of both allergen challenge and treatment with CS and saline solution.

## Discussion

This paper includes preclinical and clinical data demonstrating the safety and efficacy of a carrageenan- and sorbitol -containing (CS) nasal spray. The in vitro/ex vivo data indicate that the formulation is osmotically active while preserving the barrier-forming, virus-blocking capacity of the carrageenan. The clinical data show that the CS nasal spray is safe and well tolerable in participants with moderate to severe SAR. Although the primary endpoint based on the subjective rating of nasal congestion was not met, two objective parameters, nasal airflow and nasal secretion, showed a significant improvement upon treatment with CS nasal spray. Nasal airflow increased upon CS administration, but decreased upon administration of saline solution, leading to a significantly higher airflow in CS treated participants at the end of the challenge. The majority (60%) of participants had an increased nasal airflow after CS, but only 34% had an increased nasal airflow after SS administration. The amount of nasal secretion was reduced both after CS and SS administration, but this reduction was significant only after the CS. The low incidence of adverse events, none of them considered treatment-related, suggested safety of CS nasal spray similar to saline solution used in this study and similar to carrageenan-only (no sorbitol) nasal spray as demonstrated in previous studies.^11–14,16,24,25^

The beneficial effect of the CS nasal spray is presumable achieved via multiple modes of action attributed to carrageenan and sorbitol. First, carrageenan has excellent mucoadhesive properties that are e.g. exploited for intranasal drug delivery.^26^ We hypothesize that a mucoadhesive layer of carrageenan forms a protective barrier in the nasal mucosa that prevents small particles like pollen and dust to enter the nasal mucosa and hinders further induction or aggravation of AR symptoms like nasal congestion and nasal secretion.^17^

Secondly, polyols like sorbitol are known and widely used as humectants in the cosmetics and food industry based on their hygroscopic properties.^27^ In the context of rhinitis, xylitol, another polyol with similar properties as sorbitol, was shown to keep the nasal passages and sinuses moist and clean for a longer time than saline alone. 5-days-use of a hyperosmolar xylitol-containing nasal spray led to significant improvement of the overall quality of life score compared to pre-treatment in participants suffering from nasal obstruction.^28^ Moreover, a xylitol solution was as effective in the treatment of rhinitis medicamentosa in rats as the glucocorticoid mometasone in the reversal of histopathological changes caused by long-term treatment with oxymetazoline.^29^

Strengths of this study include the cross-over design, in which each participant serves as their own control, the random assignment to minimize possible effects from the order of treatments, and the blinding of investigators, site personnel, and the sponsor’s staff. Another strength is the use of an environmental challenge chamber to induce AR symptoms, which allows to control environmental conditions like temperature, humidity, and allergen type and concentration, and thus enables the performance of allergology studies out of allergy season and under uniform allergen exposure conditions. This limits variation and helps reducing the number of study participants. Moreover, use of the challenge chamber allows the study personnel to supervise administration of medication and documentation of outcomes, thereby enhancing participant compliance.^30–36^

The study has several limitations. One of them is the selection of the NCSS, a subjective scoring scale, as primary endpoint. The rationale for the selection of the primary endpoint was that nasal congestion comes with a significant impact upon patients’ QOL, which is considered an important determinant of the severity of nasal diseases.^37,38^ In fact, the degree of health-related QOL impairment has been demonstrated to drive patients’ choice between treatment options.^39^ Assessment of QOL in the form of patient reported outcome measures (PROMs) is regarded a standard outcome measures in clinical trials, acknowledging the fact that the classical, objective outcome variables may only partially characterize the disease of the patient. However, the focus on a PROM as primary endpoint also poses problems due to the low degree of correlation between subjective and objective outcomes assessing nasal symptoms, as systematically reviewed by Ta et al.^40^ The authors consequently recommend to use objective outcome measures to complement and confirm validated patient reported outcomes.^40^

The findings of our study support this conclusion, showing discrepancies between subjective and objective evaluations. As described in the results section, only very slight differences between groups and between timepoints were observed by NCSS that may possibly reach significance only with a much larger sample size. In contrast, differences between CS and SS in nasal airflow improvement measured by AAR became significant towards the end of the allergen challenge, indicating that this sensitive method is able to pick up subtle changes that cannot possibly be detected by PROMs like the NCSS with the available number of participants. Rhinomanometry enables the objective and accurate measurement of nasal congestion, and is considered the gold standard for measuring nasal airway patency and resistance.^41^ The method has been demonstrated to be sensitive in quantifying nasal patency after nasal provocation testing and to assess the efficacy of medications used to treat nasal congestion/obstruction.^42^ The implementation of rhinomanometry as objective endpoint in addition to the subjective symptom scores is therefore a particular upside of this study. Analogously, objective determination of nasal secretion revealed a significant reduction of nasal secretion after treatment compared to pre-treatment, which was not captured by the TNSS with sufficient sensitivity.

In this study, we used the time window from 2 to 4h after start of allergen exposure, that is, starting 15min after treatment administration and ending 2h15min after treatment administration. This interval was selected based on the expectation that the most pronounced effect of the treatment would manifest shortly after treatment. The mean residence time of carrageenan at the mucosa of approximately four hours was determined in a prior study using nasal mucociliary clearance (NMC) time assessment in healthy volunteers,^15^ and we expected the most pronounced effect to manifest in the first half of this period. However, nasal airflow continuously increased from post-treatment until the end of the allergen challenge period.

In sum, based on our findings, we propose the CS as safe and effective treatment of mild to moderate AR.

## Conclusion

Coldamaris akut, a carrageenan- and sorbitol containing nasal spray, is considered safe and effective in the relief of nasal symptoms in adults with grass pollen allergy.

### Ethics Statement

The study was conducted in Austria in accordance with in accordance with the Declaration of Helsinki on Ethical Principles for Medical Research Involving Human Subjects, the International Council for Harmonisation Guideline on Good Clinical Practice, and all applicable local regulatory requirements and laws. The study was approved by the Ethics Committee of the City of Vienna (protocol code COA_19_03, EK 19/277/1219). Informed consent was obtained from all study participants.

## Data Availability

All data produced in the present study are available upon reasonable request to the authors

## Acknowledgments

This research was sponsored by Marinomed Biotech AG.

## Disclosure

NU, MM, HD and EP are employees of Marinomed Biotech AG. MS received consulting fees from Marinomed Biotech AG. The other authors have no competing interests in this work.

This manuscript is also available at medRxiv preprint server.

## Supplementary Tables

**Table S1:** Clinical data: Nasal congestion symptom score (NCSS) of all time points for the FAS. N=40 for all timepoints.

|  | Carrageenan-Sorbitol (CS) nasal spray |  |  |  |  |  |  | Saline solution (SS) |  |  |  |  |  |
| --- | --- | --- | --- | --- | --- | --- | --- | --- | --- | --- | --- | --- | --- |
| Time | Mean | SD | LQ | UQ | Min | Max |  | Mean | SD | LQ | UQ | Min | Max |
| 00:00 | 0.1 | 0.3 | 0.0 | 0.0 | 0 | 1 |  | 0.1 | 0.3 | 0.0 | 0.0 | 0 | 1 |
| 00:15 | 0.8 | 0.5 | 0.0 | 1.0 | 0 | 2 |  | 0.7 | 0.6 | 0.0 | 1.0 | 0 | 2 |
| 00:30 | 1.4 | 0.6 | 1.0 | 2.0 | 0 | 3 |  | 1.3 | 0.6 | 1.0 | 2.0 | 0 | 2 |
| 00:45 | 1.7 | 0.7 | 1.0 | 2.0 | 0 | 3 |  | 1.6 | 0.6 | 1.0 | 2.0 | 1 | 3 |
| 01:00 | 2.0 | 0.6 | 2.0 | 2.0 | 1 | 3 |  | 1.9 | 0.6 | 2.0 | 2.0 | 1 | 3 |
| 01:15 | 2.0 | 0.7 | 2.0 | 2.0 | 0 | 3 |  | 1.9 | 0.7 | 1.0 | 2.0 | 1 | 3 |
| 01:30 | 2.2 | 0.6 | 2.0 | 3.0 | 1 | 3 |  | 2.0 | 0.6 | 2.0 | 2.0 | 1 | 3 |
| 01:45 | 2.3 | 0.7 | 2.0 | 3.0 | 1 | 3 |  | 2.2 | 0.5 | 2.0 | 2.0 | 1 | 3 |
| 02:00 | 2.1 | 0.7 | 2.0 | 2.2 | 1 | 3 |  | 1.9 | 0.6 | 1.8 | 2.0 | 1 | 3 |
| 02:15 | 2.2 | 0.7 | 2.0 | 3.0 | 1 | 3 |  | 2.0 | 0.6 | 2.0 | 2.0 | 1 | 3 |
| 02:30 | 2.1 | 0.6 | 2.0 | 3.0 | 1 | 3 |  | 2.0 | 0.7 | 2.0 | 2.2 | 1 | 3 |
| 02:45 | 2.1 | 0.7 | 2.0 | 3.0 | 1 | 3 |  | 2.0 | 0.7 | 1.8 | 3.0 | 1 | 3 |
| 03:00 | 2.2 | 0.6 | 2.0 | 3.0 | 1 | 3 |  | 2.0 | 0.7 | 2.0 | 2.2 | 1 | 3 |
| 03:15 | 2.1 | 0.7 | 2.0 | 3.0 | 1 | 3 |  | 2.0 | 0.7 | 2.0 | 3.0 | 1 | 3 |
| 03:30 | 2.2 | 0.7 | 2.0 | 3.0 | 1 | 3 |  | 2.1 | 0.7 | 2.0 | 3.0 | 1 | 3 |
| 03:45 | 2.2 | 0.7 | 2.0 | 3.0 | 1 | 3 |  | 2.1 | 0.7 | 2.0 | 3.0 | 1 | 3 |
| 04:00 | 2.2 | 0.8 | 2.0 | 3.0 | 0 | 3 |  | 2.2 | 0.7 | 2.0 | 3.0 | 1 | 3 |
| 04:15 | 2.1 | 0.8 | 2.0 | 3.0 | 0 | 3 |  | 2.1 | 0.8 | 1.8 | 3.0 | 0 | 3 |
| 04:30 | 2.2 | 0.8 | 2.0 | 3.0 | 0 | 3 |  | 2.1 | 0.7 | 2.0 | 3.0 | 1 | 3 |
| 04:45 | 2.2 | 0.7 | 2.0 | 3.0 | 1 | 3 |  | 2.2 | 0.7 | 2.0 | 3.0 | 1 | 3 |
| 05:00 | 2.2 | 0.7 | 2.0 | 3.0 | 1 | 3 |  | 2.2 | 0.7 | 2.0 | 3.0 | 1 | 3 |
| 05:15 | 2.4 | 0.7 | 2.0 | 3.0 | 1 | 3 |  | 2.2 | 0.7 | 2.0 | 3.0 | 1 | 3 |
| 05:30 | 2.2 | 0.8 | 2.0 | 3.0 | 1 | 3 |  | 2.2 | 0.7 | 2.0 | 3.0 | 1 | 3 |
| 05:45 | 2.3 | 0.7 | 2.0 | 3.0 | 1 | 3 |  | 2.2 | 0.7 | 2.0 | 3.0 | 1 | 3 |
| 06:00 | 2.3 | 0.8 | 2.0 | 3.0 | 1 | 3 |  | 2.2 | 0.7 | 2.0 | 3.0 | 1 | 3 |

Abbreviations: CS, carrageenan-sorbitol containing nasal spray. SS, saline solution; SD, standard deviation; LQ, lower quartile; UQ, upper quartile.

**Table S2:** Clinical data: Mean difference in NCSS [Pre-treatment - ø(2-4h)] for the FAS.

|  | **CS** | **SS** | **CS – SS** |
| --- | --- | --- | --- |
| Mean | 0.16 | 0.11 | 0.02 |
| SD | 0.50 | 0.53 | 0.66 |
| Median | 0.00 | 0.00 | 0.11 |
| Min | -0.89 | -0.78 | -1.56 |
| Max | 1.44 | 1.56 | 1.33 |
| N | 40<sup>1</sup> | 40<sup>1</sup> | 39<sup>2</sup> |

Abbreviations: NCSS, Nasal Congestion Symptom Score. CS, carrageenan-sorbitol containing nasal spray. SS, saline solution.
SD, standard deviation. Min, minimum value. Max, maximum value.
1 n = 40 out of 41 patients completed each treatment period
2 n = 39 out of 41 patients completed both treatment periods

**Table S3:**
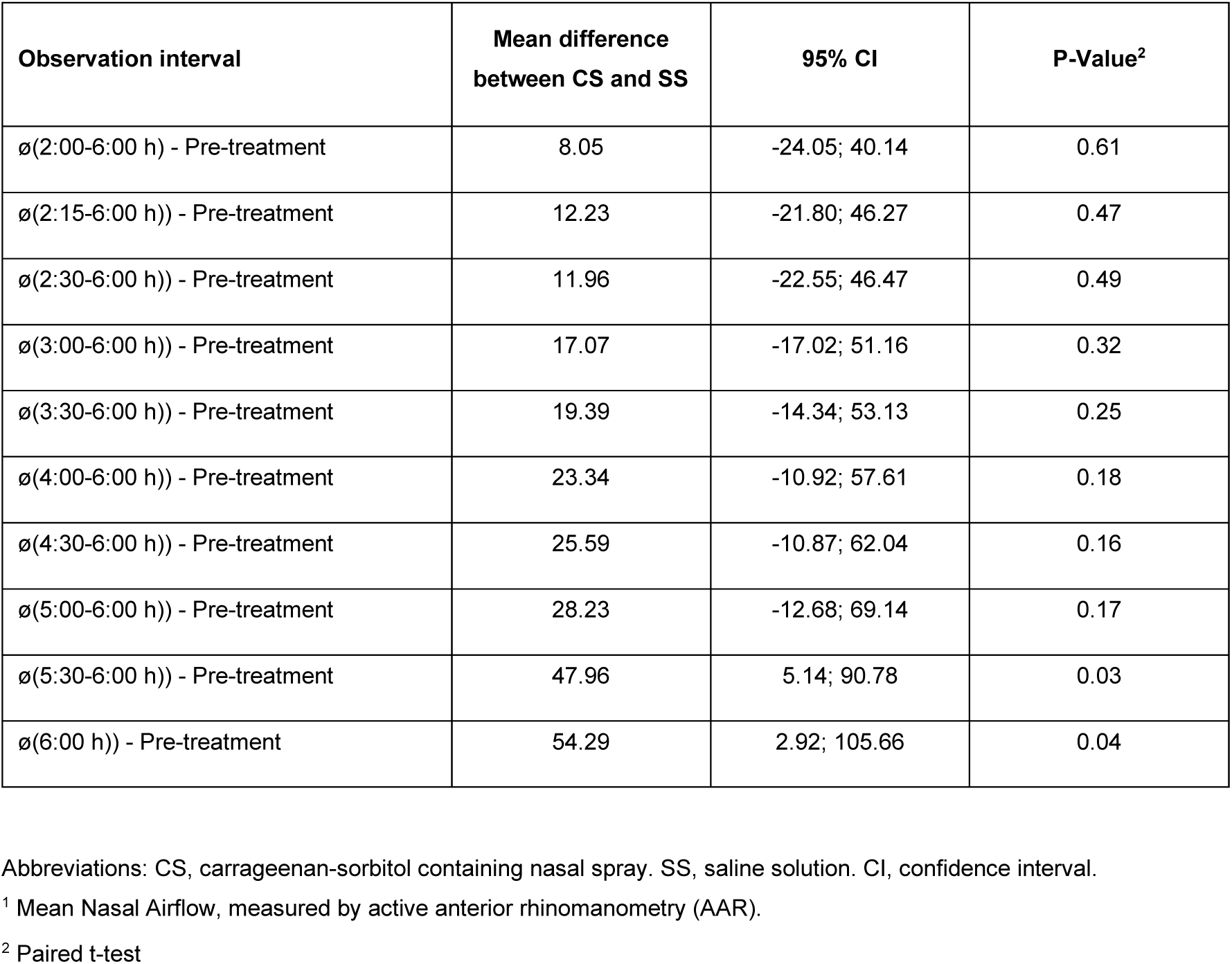
Clinical data: Mean difference in AAR change from pre- to post-treatment between CS and SS for the FAS. Values for the respective treatment period are first calculated individually by subtracting the mean pre-treatment airflow from the mean nasal airflow over the indicated post-treatment time period. Differences between treatments are computed likewise by subtracting the [mean pre- to post-treatment difference for SS] from the [mean pre- to post-treatment difference for CS]. Paired t-tests were applied to those differences. Differences above 0 are favorable for the CS treatment.

| Observation interval | Mean difference between CS and SS | 95% CI | P-Value<sup>2</sup> |
| --- | --- | --- | --- |
| ø(2:00-6:00 h) - Pre-treatment | 8.05 | -24.05; 40.14 | 0.61 |
| ø(2:15-6:00 h)) - Pre-treatment | 12.23 | -21.80; 46.27 | 0.47 |
| ø(2:30-6:00 h)) - Pre-treatment | 11.96 | -22.55; 46.47 | 0.49 |
| ø(3:00-6:00 h)) - Pre-treatment | 17.07 | -17.02; 51.16 | 0.32 |
| ø(3:30-6:00 h)) - Pre-treatment | 19.39 | -14.34; 53.13 | 0.25 |
| ø(4:00-6:00 h)) - Pre-treatment | 23.34 | -10.92; 57.61 | 0.18 |
| ø(4:30-6:00 h)) - Pre-treatment | 25.59 | -10.87; 62.04 | 0.16 |
| ø(5:00-6:00 h)) - Pre-treatment | 28.23 | -12.68; 69.14 | 0.17 |
| ø(5:30-6:00 h)) - Pre-treatment | 47.96 | 5.14; 90.78 | 0.03 |
| ø(6:00 h)) - Pre-treatment | 54.29 | 2.92; 105.66 | 0.04 |

Abbreviations: CS, carrageenan-sorbitol containing nasal spray. SS, saline solution. CI, confidence interval.
<sup>1</sup> Mean Nasal Airflow, measured by active anterior rhinomanometry (AAR).
<sup>2</sup> Paired t-test

**Supplemental Figure S1:**
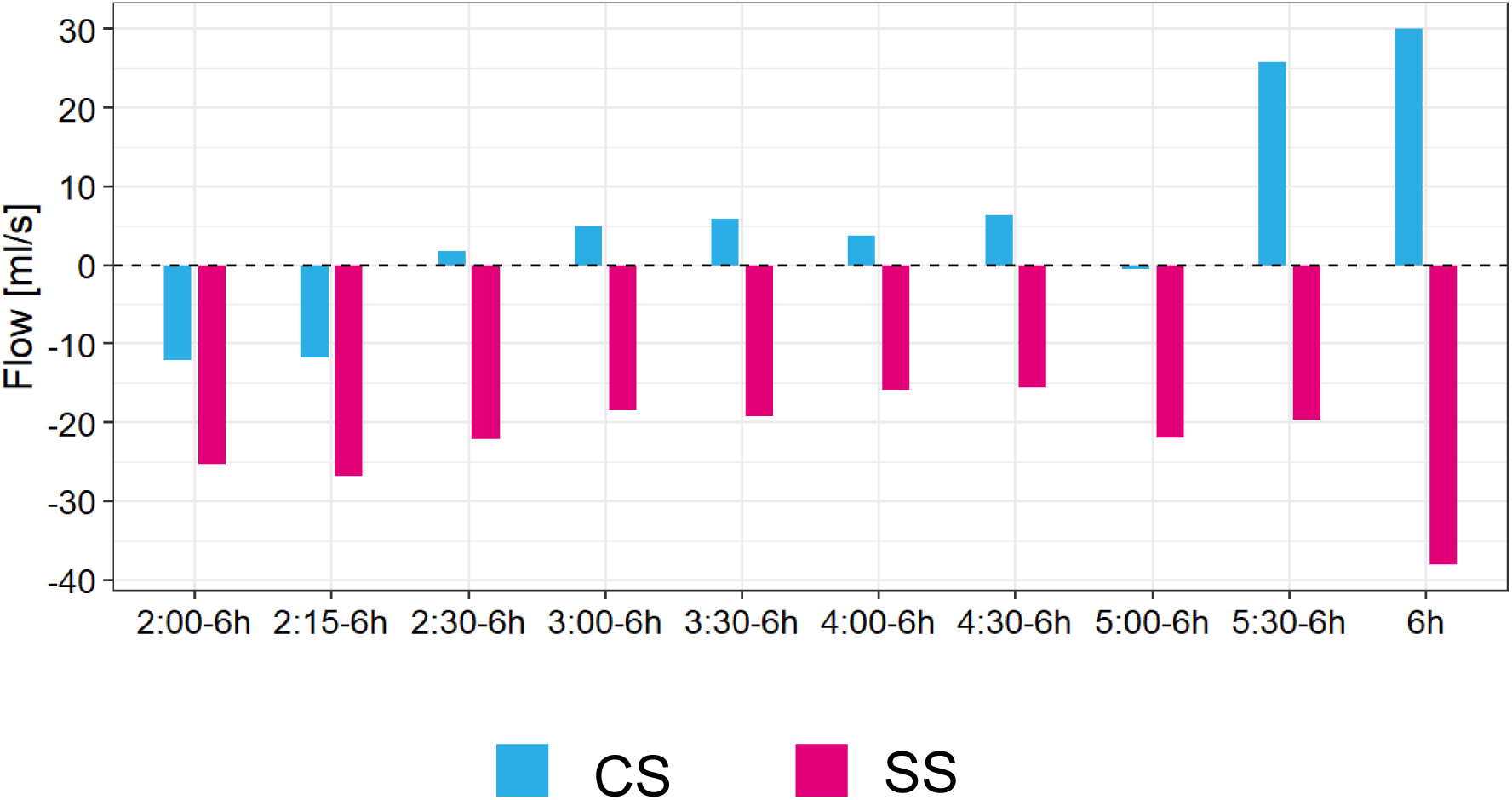
Clinical data: Median anterior nasal airflow absolute differences to pre-treatment for the FAS. Differences in post-treatment nasal airflow compared to pre-treatment in the CS group (cyan) and the SS (magenta). Positive values indicate higher nasal airflow post-treatment, negative value indicate lower nasal airflow post-treatment compared to pre-treatment.

